# Strength of spatial correlation between structural brain network connectivity and brain-wide patterns of proto-oncogene and neural network construction gene expression is associated with diffuse glioma survival

**DOI:** 10.1101/2023.11.27.23299085

**Authors:** Shelli R. Kesler, Rebecca A. Harrison, Alexa De La Torre Schultz, Hayley Michener, Paris Bean, Veronica Vallone, Sarah Prinsloo

## Abstract

Like other forms of neuropathology, gliomas appear to spread along neural pathways. Accordingly, our group and others have previously shown that brain network connectivity is highly predictive of glioma survival. In this study, we aimed to examine the molecular mechanisms of this relationship via imaging transcriptomics. We retrospectively obtained presurgical, T1-weighted MRI datasets from 669 adult patients, newly diagnosed with diffuse glioma. We measured brain connectivity using gray matter networks and coregistered these data with a transcriptomic brain atlas to determine the spatial co-localization between brain connectivity and expression patterns for 14 proto-oncogenes and 3 neural network construction genes. We found that all 17 genes were significantly co-localized with brain connectivity (p < 0.03, corrected). The strength of co-localization was highly predictive of overall survival in a cross-validated Cox Proportional Hazards model (mean area under the curve, AUC = 0.68 +/- 0.01) and significantly (p < 0.001) more so for a random forest survival model (mean AUC = 0.97 +/- 0.06). Bayesian network analysis demonstrated direct and indirect causal relationships among gene-brain co-localizations and survival. Gene ontology analysis showed that metabolic processes were overexpressed when spatial co-localization between brain connectivity and gene transcription was highest (p < 0.001). Drug-gene interaction analysis identified 84 potential candidate therapies based on our findings. Our findings provide novel insights regarding how gene-brain connectivity interactions may affect glioma survival.

## Introduction

Diffuse gliomas are malignant neoplasms of the brain that have been shown to provoke whole-brain network disruption regardless of their primary foci (1–3). It is well-known that neuropathology propagates throughout the brain from the initial sites of neurodegeneration via mechanisms such as transneuronal spread (4) and neuron derived exosomes (5), for example. In other words, pathology uses the same neural pathways and mechanisms that information uses, and the more active or connected these pathways are, the more pathology will be transmitted. In the case of glioma, tumors form and reorganize neural networks and glioma-infiltrated neurons show significant hyperactivity. The more connected these neurons are, the more efficiently the tumor proliferates to other areas of the brain, and the shorter the patient’s survival (6, 7).

Accordingly, previous studies have demonstrated that whole-brain network connectivity is highly predictive of glioma survival (8, 9). Uniquely, our prior work has shown that brain-wide gray matter connectivity accurately predicts glioma genotype and overall survival, performing significantly better than demographic, clinical, or tumor radiomic models (10, 11). Gray matter volumes extracted from T1-weighted brain MRI (T1MRI) demonstrate covariance patterns that reflect structural and functional connectivity networks (12, 13). T1MRI is non-invasive, efficient to acquire, provides excellent contrast between gray and white matter, and tends to be standard of care for glioma patients. However, T1MRI does not measure molecular mechanisms. Fortunately, advances in the development of transcriptomic brain atlases make it possible to co- localize neuroimaging and gene expression data (14). Known as imaging transcriptomics, this emerging field has already provided novel insights regarding the biological pathways underlying brain health and disease (15).

Krishna et al. (2023) performed RNA sequencing of glioma tissue with high and low functional connectivity, sampled during neurosurgery (6). However, very few studies have applied the non-invasive technique of imaging transcriptomics to examine gene expression patterns in glioma. Mandal et al. (2020) co-localized brain-wide connectomics with spatial profiles of several proto-oncogenes to reveal novel information regarding regional vulnerability to glioma (16). Otherwise, Germann and colleagues (2022) published a review describing connectomics and imaging transcriptomics and their potential applications for neuro-oncology (17). Here we report the first study to investigate the relationship between brain-wide connectivity, brain-wide gene expression, and glioma survival.

## Methods

### Participant Datasets

We retrospectively identified adult (age 18 or older) patients with histopathologically confirmed World Health Organization grade II–IV gliomas who were newly diagnosed and had not yet undergone any treatment (biopsy, resection, chemoradiation, etc.). A total of 669 patients met these criteria and had an available, usable, pre-surgical, 3 tesla T1MRI. Patients were treated during the years of 1990–2022. De-identified T1MRI, demographic, and other clinical data were extracted from the electronic medical record. The University of Texas MD Anderson Cancer Center Institutional Review Board gave ethical approval of this work (protocol# 2021-0236), which included a waiver of informed consent.

### Structural Brain Connectivity Maps

Gray matter volumes were segmented from T1MRI with Voxel-Based Morphometry in Statistical Parametric Mapping v12 (Wellcome Trust Centre for Neuroimaging, London, UK) in Matlab v2023b (Mathworks, Inc., Natick, MA). We employed Diffeomorphic Anatomical Registration Through Exponentiated Lie Algebra (DARTEL), which uses a large deformation framework to preserve topology and employs customized, sample-specific templates resulting in superior image registration, even in lesioned brains, compared to other automated methods (18). Successful normalization was confirmed using visual and quantitative quality assurance methods.

A gray matter covariance map was constructed for each participant using a similarity- based extraction method (19). Specifically, network nodes were defined as 3×3×3mm cubes spanning the entire gray matter volume (i.e., 54 gray matter values per cube, 7134 +/- 382 cubes). A correlation matrix was calculated across all pairs of nodes. We then applied graph theoretical analysis using the bNets Toolbox v2.2 (Brain Health Neuroscience Lab, Austin, TX) (20) and Brain Connectivity Toolbox v2019-03-03 (21) to calculate local efficiency (22) for each node. Nodal efficiency is consistently observed to be affected in patients with diffuse glioma (2, 23–25) and we have shown that it predicts overall survival and IDH tumor status with cross-validated areas under the curve of 0.88 and 0.94, respectively (10, 11). A statistical image corresponding to the nodal efficiency of each cube was created using the standard space Montreal Neurological Institute (MNI) coordinates of each cube.

### Gene Expression Profiles

We obtained brain transcriptome data from the Allen Human Brain Atlas (AHBA), which is currently considered the most comprehensive transcriptional brain map available (15). AHBA was developed using six adult human donor brains to provide expression data from tens of thousands of genes measured from thousands of brain regions (26). The standard space T1MRI’s of the donor brains are also publicly available to facilitate their spatial coregistration with neuroimaging data from study samples. Mandal and colleagues showed that several proto-oncogenes important for gliomagenesis, as identified in a review by Molinaro et al. (27), were spatially correlated with glioma distribution (16). Krishna and colleagues showed that expression of certain genes involved in the development of neural networks were upregulated in highly connected, tumor-infiltrated brain regions (6). We therefore extracted expression profiles for these same genes, excluding those for which there were no valid AHBA probes available (Supplementary Table 1).

### Imaging Transcriptomics

To determine the co-localization of brain imaging and transcription data, we used the Multimodal Environment for Neuroimaging and Genomic Analysis (MENGA) toolbox v3.1 in Matlab v2023b to spatially correlate the structural brain connectivity map of each participant with patterns of AHBA expression from the 17 selected genes. The details of this analysis were described by Rizzo et al. (2016) (28). Briefly, the imaging data for each glioma participant was resampled into AHBA coordinates with a 3mm resolution. The expression data for each AHBA donor, for each gene was sampled from each of 169 AHBA regions resulting in a 169×6 region-by-gene expression matrix for each gene. Principal Components Analysis was performed on the region-by-gene expression matrix to identify components explaining at least 95% variance across the AHBA donors. The component scores were then entered as independent variables into a weighted, multiple linear regression analysis with the corresponding local efficiency values from the glioma participant as the dependent variable. The regression weights were the mean number of samples in each connectivity map region and the variability of these connectivity values. P values for the regression models were corrected for multiple comparisons using false discovery rate (29).

### Association of Imaging Transcriptomics with Survival

The regression R squared statistics for each gene for each participant (669×17 matrix) were then entered into 1) Cox Proportional Hazards (PH), and 2) random forest survival (30) regression models to predict overall survival in months. The absolute value of the R squared statistics were utilized as predictors to determine if the strength of the image-transcriptome spatial co-localization was significantly associated with survival. Compared to inferential models like Cox PH, random forest models tend to demonstrate significantly better performance, are more robust to multicollinearity, and can capture complex non-linear relationships and interactions between features. However, they have lower interpretability as the importance of individual predictors is more difficult to ascertain. There is also increased risk of overfitting the model to the data.

We implemented 10-folds cross-validation (31) for both models to reduce overfitting and increase external validity. Specifically, the N=669 cases were randomly shuffled and then split into 10 subsets (i.e., folds). For each of the 10 cross-validation loops, a Cox PH/random forest model was trained on the data from 9 of the folds and then tested on the left-out fold such that every fold was tested once. The time-dependent area under the curve (AUC) of the receiver operating characteristic was used to determine model performance. This is the integral of AUC on the range of survival time from 0 to maximum, weighted by the estimated probability density of the time-to-event outcome. This AUC calculation accounts for censoring and the time-dependent nature of the parameters (32). The AUC was averaged across the 10 cross- validation loops for each model and the means were compared using a t-test. For the cross-validation models with the highest AUC, we examined the coefficients and p values (Cox PH) and the variable holdout error (random forest survival) (33) to interpret the importance of individual genes in the models.

### Path Analysis

We conducted exploratory Bayesian network analysis to examine the causal relationship among the genes whose spatial co-localization with gray matter connectivity showed significant association with survival across both predictive models. Unlike Structural Equation Modeling, this method learns the conditional dependencies between variables without any prior assumptions. We utilized a hill-climbing approach by optimizing the Bayesian information criterion to identify the best fit network. The conditional independence/dependence relationships among variables were encoded with a directed acyclic graph. We employed the method described by Stajduhar and Dalbelo-Basic (34) to account for censored data.

### Reliability of Genomic Data

Most of the genes included in AHBA have multiple probes with some showing more reliable expression patterns than others. First, the current accuracy of the probe to gene mapping was verified and probe data were normalized to z-scores. Representative probes were then selected in a data-driven manner considering between-donor homogeneity and the distributions of probe data. Genomic autocorrelations were calculated to measure the gene expression variability between donors. Most of the tissue samples used to develop AHBA were obtained only from the left hemisphere and therefore, we employed a binary mask to limit our analysis to the left hemisphere (28).

### Gene Enrichment

We performed a gene ontology enrichment analysis using the 17 selected genes ranked by the R squared statistic from the co-localization analysis. We utilized the Gene Ontology Enrichment Analysis and Visualization (GOrilla) online tool, which identifies hierarchical gene ontology terms by optimizing the hypergeometric tail probability across subsets of genes based on ranking (35, 36).

### Drug-Gene Interactions

We utilized the web-based Drug-Gene Interaction Database (DGIdb) v5.0 (37) to explore the potential druggability of genes whose spatial co-localization with gray matter connectivity showed significant association with survival across both predictive models. DGIdg identifies drug-gene interactions based on publications and other open sources and calculates a Query Score for ranking search results. The Query Score considers the specificity of the gene-drug association, the overlapping drug interactions among all genes in the query, and the number of sources supporting the interaction. DGIdb results also describe the indication for drugs, when known, as well as the current regulatory status.

## Results

### Patient Characteristics

Patients were aged 48 +/- 16 years (range: 18 to 87) at diagnosis and 59% were male. Most patients had high grade tumors (65%) and underwent gross total resection (46%). Left hemisphere (75%) and frontal lobe (48%) were the most common tumor locations, and most were histologic astrocytomas (76%). Those patients with available tumor genotyping data were relatively split between IDH mutant and wild type (N = 91 missing) as well as MGMT methylated and unmethylated (N = 450 missing). Most patients (98%) had a Karnofsky Performance Scale (KPS) of 70 or higher, although these data were missing for over half the cohort (Table 1). Median overall survival was 81 months (95% CI: 70 to 96, Figure 1a) with a 36.5% mortality rate over the follow up period of 335 months (median = 45 months).

**Figure 1.**
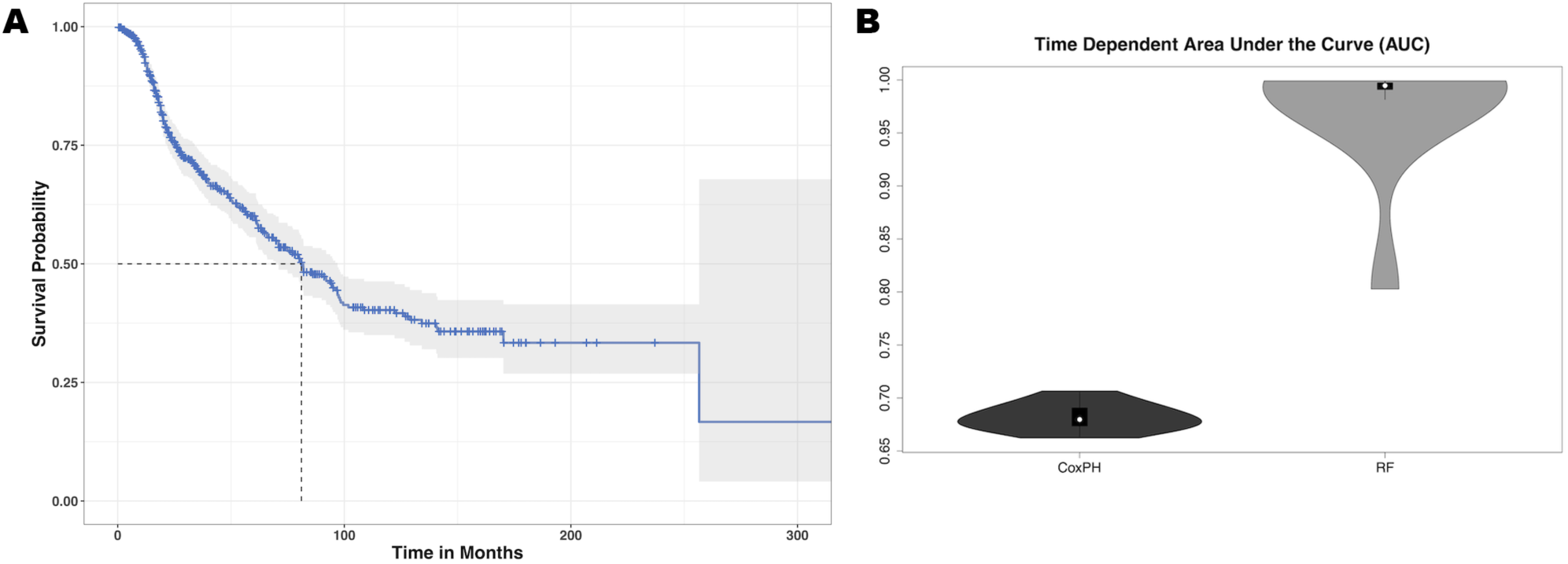
Glioma Survival Data. A: Median overall survival for our cohort of glioma patients was 81 months. B: The mean cross-validated, time dependent area under the curve for predicting survival from imaging transcriptomic relationships was 0.68 +/- 0.01 for Cox PH and 0.97 +/- 0.06 for random forest (RF). Random forest significantly outperformed Cox PH (p < 0.001, corrected).

**Table 1.**
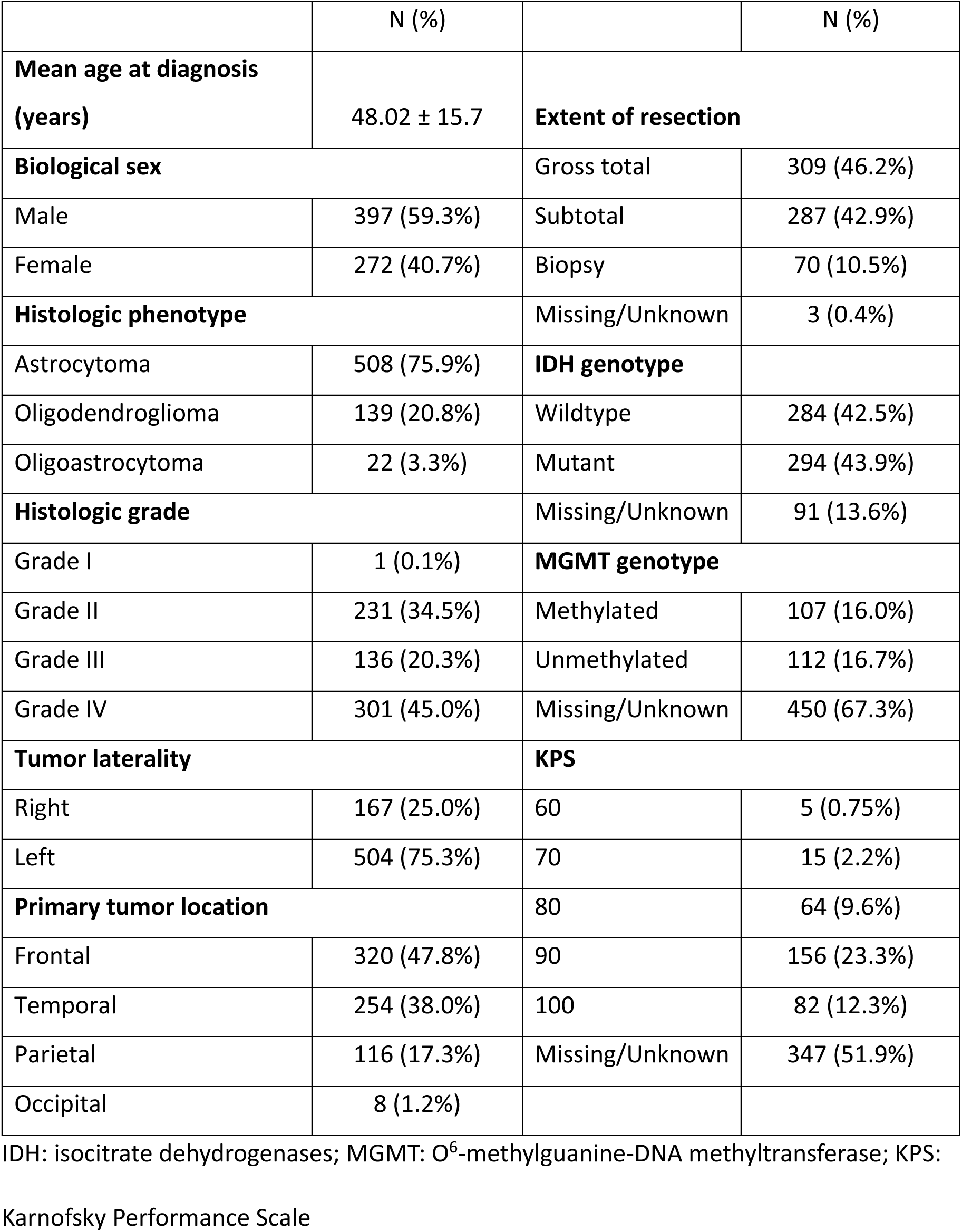
Patient Characteristics.

### Imaging Transcriptomics

As shown in Table 2, all 17 selected genes demonstrated significant spatial co-localization with whole brain gray matter connectivity, even after correction for multiple comparisons (p < 0.03, corrected).

**Table 2.** Imaging Transcriptomics. Multiple regression analysis indicated significant spatial co- localization of gene expression and gray matter connectivity. FDR: false discovery rate, sd: standard deviation.

| Gene | R<sup>2</sup> | F | p<br>uncorrected | p FDR<br>corrected | Direction of<br>relationship | Genomic<br>autocorrelation<br>coefficient [R<sup>2</sup><br>(sd)] |
| --- | --- | --- | --- | --- | --- | --- |
| IDH1 | 0.335 | 14.213 | <0.001 | <0.001 | -1 | 0.16 (0.14) |
| IDH2 | 0.058 | 5.189 | 0.002 | 0.003 | 1 | 0.39 (0.15) |
| TERT | 0.227 | 9.292 | <0.001 | <0.001 | -1 | 0.24 (0.10) |
| ATRX | 0.311 | 10.240 | <0.001 | <0.001 | 1 | 0.14 (0.07) |
| EGFR | 0.219 | 8.973 | <0.001 | <0.001 | -1 | 0.28 (0.11) |
| CDKN2A | 0.004 | 3.139 | 0.030 | 0.030 | -1 | 0.54 (0.09) |
| CDKN2B | 0.121 | 5.619 | <0.001 | 0.001 | -1 | 0.15 (0.14) |
| PTEN | 0.305 | 12.660 | <0.001 | <0.001 | 1 | 0.10 (0.10) |
| TP53 | 0.205 | 6.682 | <0.001 | <0.001 | -1 | 0.02 (0.03) |
| NF1 | 0.267 | 8.662 | <0.001 | <0.001 | -1 | 0.05 (0.05) |
| MDM2 | 0.286 | 9.325 | <0.001 | <0.001 | -1 | 0.09 (0.07) |
| PIK3CA | 0.272 | 11.154 | <0.001 | <0.001 | -1 | 0.16 (0.09) |
| FUBP1 | 0.144 | 5.006 | <0.001 | 0.001 | 1 | 0.13 (0.10) |
| NOTCH1 | 0.246 | 10.045 | <0.001 | <0.001 | -1 | 0.20 (0.14) |
| **SYNPO** | 0.069 | 4.137 | 0.004 | 0.004 | 1 | 0.50 (0.11) |
| **NTNG1** | 0.068 | 17.063 | <0.001 | <0.001 | -1 | 0.66 (0.09) |
| **THBS1** | 0.211 | 6.858 | <0.001 | <0.001 | 1 | 0.04 (0.06) |

### Imaging Transcriptome and Survival

The mean cross-validated, time dependent AUCs were 0.68 +/- 0.01 for Cox PH and 0.97 +/- 0.06 for random forest (Figure 1b). Random forest significantly outperformed Cox PH (p < 0.001, corrected). In the best Cox PH model (AUC = 0.71), co-localization of brain connectivity and *TERT* (*B* = -5.1, p = 0.004), *ATRX* (*B* = -5.5, p = 0.012), *FUBP1* (*B* = 5.2, p = 0.03), *NOTCH1* (*B* = -7.5, p = 0.004), and *CDKN2A* (*B* = 1.0, p = 0.02) were significant predictors (Table 3). In the best random forest model (AUC = 0.999), the most important predictors were the strengths of co-localization of brain connectivity with *TERT, ATRX, FUBP1*, and *SYNPO* (Table 4). Figure 2 shows the causal, Bayesian network pathways among spatial co-localization and survival.

**Figure 2.**
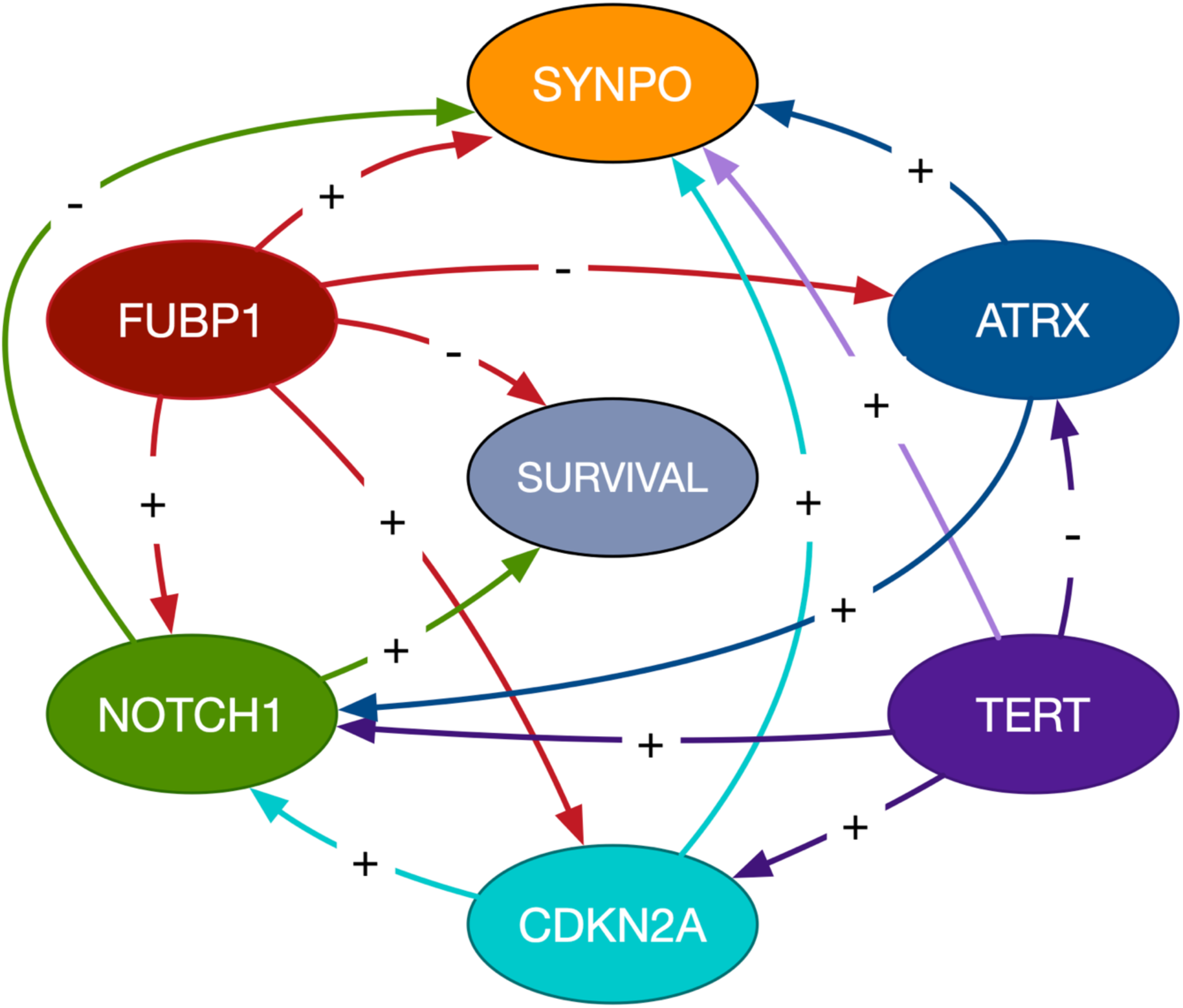
Bayesian Network Path Analysis. Causal relationships are represented by directed arrows between imaging transcriptomes and glioma survival. The gene labeled nodes represent the *strength of the relationship between the gene and brain connectivity*, not gene expression itself. The direction of the coefficient is represented by a + for direct and - for inverse.

**Table 3.** Variable importance data from the best cross-validated Cox PH model. Variables with a p value less than 0.05 were considered the most important (bold font).

| gene | coef | z | p |
| --- | --- | --- | --- |
| **TERT** | -5.103 | -2.884 | 0.004 |
| **NOTCH1** | -7.499 | -2.870 | 0.004 |
| **ATRX** | -5.477 | -2.477 | 0.013 |
| **CDKN2A** | 10.390 | 2.415 | 0.016 |
| **FUBP1** | 5.217 | 2.115 | 0.034 |
| MDM2 | 3.938 | 1.749 | 0.080 |
| PTEN | 5.264 | 1.716 | 0.086 |
| CDKN2B | 4.641 | 1.669 | 0.095 |
| TP53 | 3.577 | 1.327 | 0.184 |
| IDH2 | 2.229 | 0.831 | 0.406 |
| EGFR | -1.685 | -0.672 | 0.502 |
| NF1 | -1.460 | -0.459 | 0.646 |
| PIK3CA | -1.097 | -0.403 | 0.687 |
| IDH1 | 0.870 | 0.300 | 0.764 |
| NTNG1 | 0.710 | 0.294 | 0.769 |
| SYNPO | 0.286 | 0.089 | 0.929 |
| THBS1 | -0.035 | -0.014 | 0.989 |

**Table 4.**
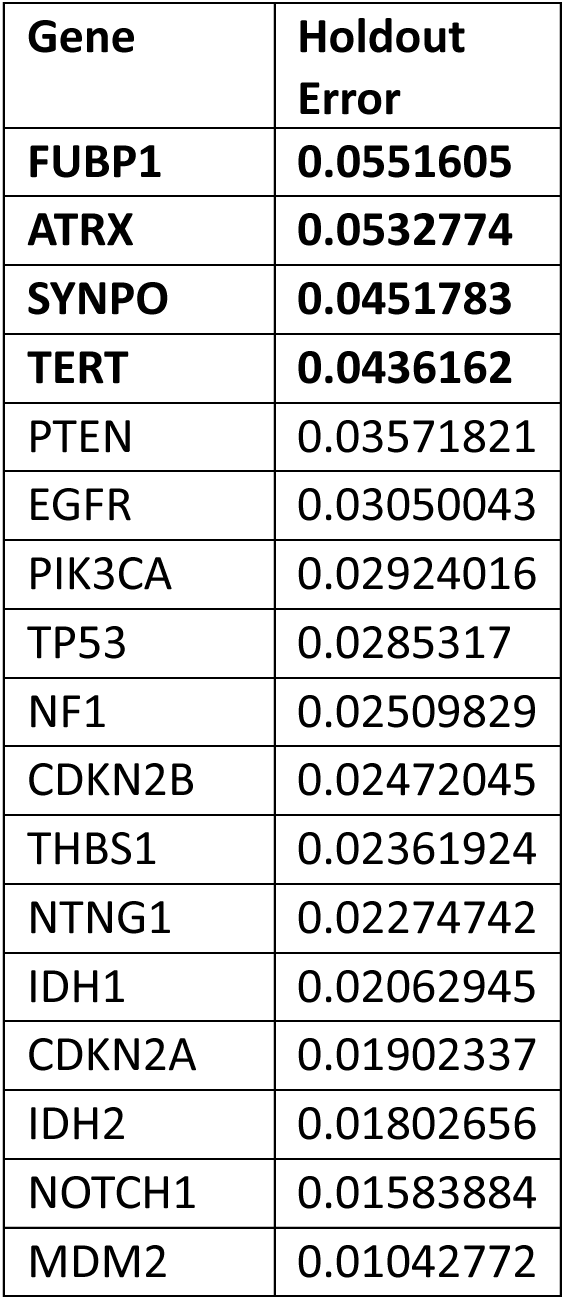
Variable importance data from the best cross-validated random forest model. The holdout error is the error of the permuted model when that variable was left out and thus, the higher the value (error), the more important the variable for the model. Variables with holdout error greater than 1 standard deviation above the mean were considered the most important (bold font).

### Gene Enrichment

Ontology analysis indicated several processes that are overexpressed when spatial co-localization between gray matter connectivity and gene transcription is highest. These included biological processes related to regulation of biological quality as well as cellular, primary, nitrogen compound, organic substance, and macromolecule metabolism (Figure 3a).

**Figure 3.**
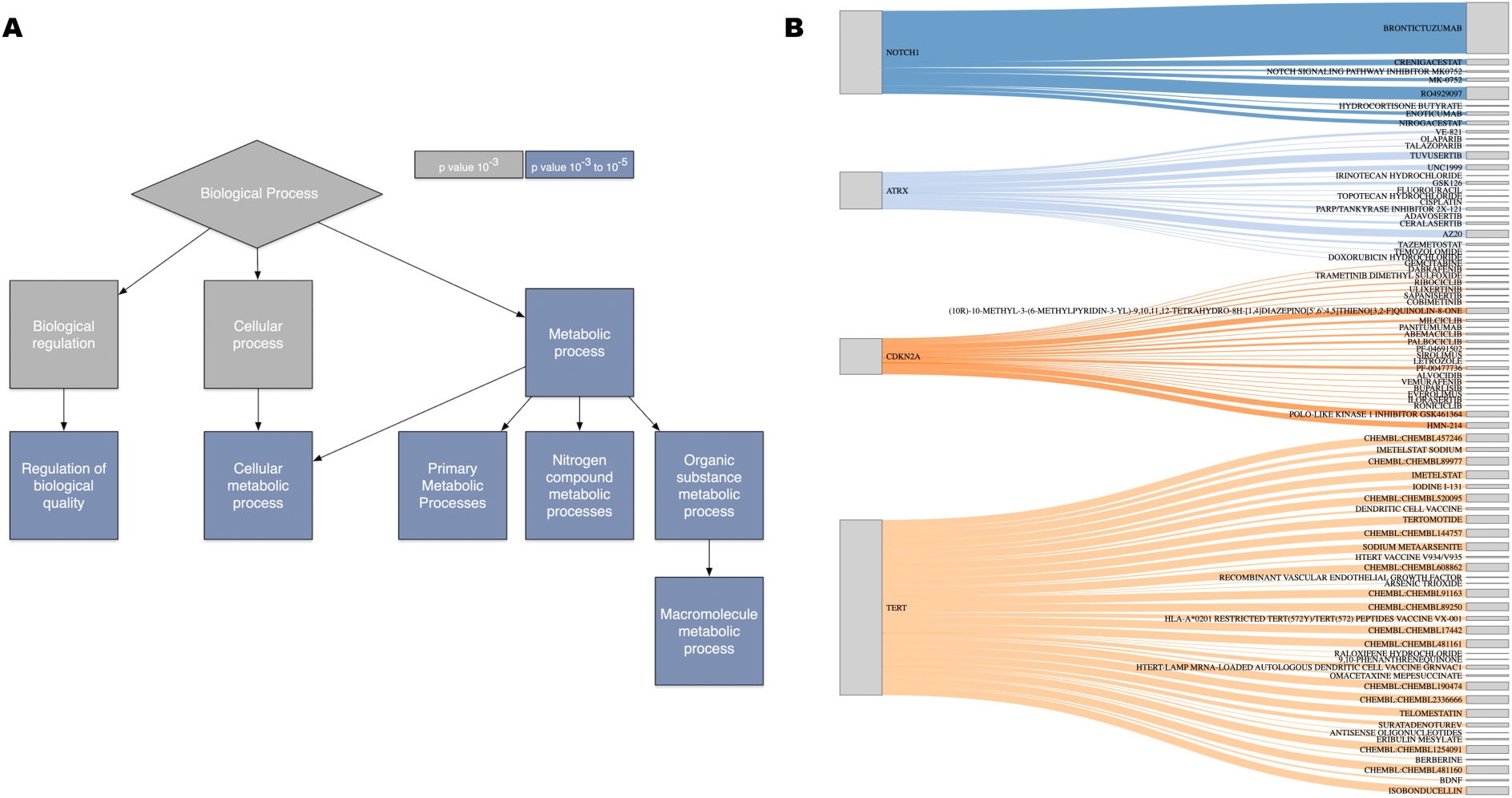
Gene Ontology and Drug-Gene Interactions. A: Gene ontology analysis of the genes whose relationship with brain connectivity was predictive of glioma survival. B: Drug-gene interaction results for these same genes. The magnitude of the Query Score is represented in the height of the gray box to the right of the drug name.

### Drug-Gene Interactions

We submitted the genes whose co-localization with brain connectivity was determined as most important across the two predictive models (*TERT, ATRX, FUBP1, CDKN2A, SYNPO* and *NOTCH1*) to the Drug-Gene Interaction Database. The database identified 84 different drugs with a mean query score of 1.7 +/- 2.6 (range = 0.02 to 22.1). The highest Query Score was for Brontictuzumab (OMP-52M51). However, there were no interactions found in the database for *FUBP1* or *SYNPO* (Figure 3b, Supplementary Table 2).

## Discussion

We conducted the first imaging transcriptomics study evaluating the relationship between widespread brain connectivity, brain gene expression, and glioma survival. We found that the expression patterns of important proto-oncogenes as well as neural network construction genes were significantly spatially correlated with whole brain, gray matter connectivity. Additionally, we showed that the strength of this co-localization was highly predictive of overall survival. Our findings make a novel contribution to results from prior studies (6, 16) by combining both proto-oncogenes and neural network construction genes and by examining large scale brain connectivity rather than tumor specific connectivity.

We evaluated brain connectivity using a metric known as nodal efficiency, which indicates how many direct connections a brain region has with other regions. Information processing is more efficient when there are direct connections among neural communities. Given previous findings that highly connected, tumor-infiltrated neurons tend to increase proliferation of glioma pathology (6), we expected higher nodal efficiency to be associated with lower survival. However, this was dependent on the gene-brain pathway. Higher expression of *ATRX, CDKN2A, FUBP1*, and *SYNPO* as well as lower expression of *TERT* and *NOTCH1* were associated with higher nodal efficiency. The strength of these relationships was significantly associated with survival. This is consistent with our prior findings that overall survival in patients with glioma is associated with areas of lower as well as areas of higher gray matter connectivity (10).

It is important to remember that these relationships are not directly between gene expression and survival, but instead reflect the effects of spatial co-localization of gene expression and brain connectivity on survival. The causal, Bayesian network path analysis provides further insight regarding these complex relationships. *NOTCH1*- and *FUBP1*-related brain connectivity showed the only direct influences on survival. *TERT*-related brain connectivity showed an indirect effect on survival by exerting influence on *CDKN2A*-related brain connectivity, which in turn influenced *NOTCH1*-related brain connectivity. *FUBP1*-related brain connectivity also appeared to influence the relationship of brain connectivity with *CDKN2A* and *NOTCH1* as well as having a direct effect on survival status. *ATRX*-related brain connectivity showed an indirect effect on survival via the relationship between *NOTCH1* and brain connectivity.

Interpreting the impact of higher versus lower brain connectivity on survival is challenging. These relationships were evaluated in multivariate space and the significantly better performance of the random forest model compared to the Cox PH model indicates that these relationships were highly nonlinear. These findings are consistent with evidence that the brain network operates as a system at criticality, functioning optimally when finely balanced on the edge of various competing demands (38). In other words, there is an optimal range of brain connectivity where too little or too much can be detrimental. Gene-interaction networks also operate at criticality (39). At the edge of criticality, these systems are highly sensitive to perturbations. This sensitivity can be beneficial, as it allows the network to flexibly respond to changing environments or conditions. However, it can also make the system vulnerable to mutations or detrimental stressors. The interactions within critical systems can lead to emergent behaviors that are not predictable by studying individual components (genes, brain regions) in isolation.

The optimal level of brain connectivity allows for the efficient exchange of information without overwhelming the system’s resources (22) and thus connectivity is inherently tied to metabolism (40). Our gene ontology analysis indicated that metabolism was the common function across the genes that were most important in our predictive models. This is consistent with previous research on cancer metabolism (41). Our Bayesian network path analysis showed that *SYNPO*-related brain connectivity was directed by the relationships between brain connectivity and all other genes in the model. The direct or indirect effects of *SYNPO*-related brain connectivity on glioma survival was not apparent from our findings. *SYNPO* expression is believed to be involved in neural network construction (6) and thus would rely heavily on upstream, metabolically driven pathways. However, the effects of *SYNPO*-related brain connectivity on survival in the context of proto-oncogenes requires further investigation.

Given that neural connectivity is associated with neural activity, it would be interesting to see if these properties could be safely reduced to slow tumor progression using drugs that impact the gene-brain connectivity relationships identified by our models. *NOTCH1-* and *FUBP1-*related connectivity showed the only direct causal links to survival and are therefore strong candidate targets. Our drug-gene interaction analysis identified Brontictuzumab (OMP- 52M51) as the best candidate. Brontictuzumab is a human monoclonal antibody that blocks *NOTCH1* signaling and has already been proposed as a potential treatment for glioma and other cancers (42). Our analysis did not identify any drug-gene interactions specific to *FUBP1*, however, this analysis suggests drugs in the context of all queried genes. We noted that the Query Score doubled from 10.91 when *NOTCH1* was entered alone to 22.11 when including all 6 significant genes. This suggests Brontictuzumab has overlapping interactions with the other genes in the query, including *FUBP1*. Our Bayesian network analysis suggested that enhancing the relationship between *NOTCH1* and brain connectivity has survival benefit. Given the nonlinear nature of these relationships, it is possible that inhibiting *NOTCH1* could enhance this relationship within a certain window of parameters. However, further research is required especially given that this analysis required query of the genes themselves. Thus, the results may not reflect candidate drugs that would directly impact the gene-brain connectivity *relationship*.

Nonpharmacologic alternatives for reducing neural activity include mindfulness meditation and neuromodulation. Mindfulness meditation focuses on decreasing intrusive thought processes and has been shown to reduce neural activity, especially in the default mode network (43), which is the most metabolically active functional brain network (40). Meditation has additional benefits such as reducing distress, inflammation, and blood pressure, and improving cognitive function (44). However, mindfulness meditation can also increase neural activity and connectivity in some individuals, especially among novices (43). Neuromodulation involves regulating brain activity via neurostimulation or neurofeedback. We previously showed that healthy adults can be trained to significantly down-regulate neural activity and improve cognitive function in only 1.5 total hours of neurofeedback across 2 weeks (45). In addition to regulating neural activity, cognitive function is itself an important predictor of survival in glioma (46). However, given the complex, nonlinear nature of our findings, it will be critical to determine which specific neural communities require up regulation and which require down regulation.

In conclusion, we demonstrated that the expression patterns of several proto-oncogenes as well as neural network construction genes are significantly co-localized with brain connectivity. However, higher brain connectivity may not always be associated with lower survival. We also showed that some gene-brain relationships have a direct effect on survival, but most others tend to influence mortality by affecting other gene-brain relationships. The limitations of our study include the retrospective nature of the data. We selected genes and measured local efficiency based on prior literature, but other genes, connectivity metrics or connectivity methods may have produced different results. As noted above, the AHBA provides limited data for the right hemisphere and contralateral differences in gene-brain relationships cannot be ruled out. There was heterogeneity in gene expression across the AHBA donors as reflected in the genomic autocorrelations, and this may have reduced the power to detect certain effects. Bayesian network coefficients are analogous to those derived from multiple regression and therefore do not adequately reflect the nonlinear relationships among variables. Despite these limitations, our findings provide novel insights regarding how gene-brain connectivity interactions may affect therapeutic vulnerabilities and glioma survival.

## Acknowledgements

This research was supported by the National Institutes of Health (1R03CA241862). The sponsor was not involved in any aspects of study design, analysis, manuscript preparation or submission.

## Author contributions

Conceptualization: SRK, RAH, SP. Data collection: HM, PB, VV, ADS, SP. Formal analysis: SRK, ADS. Methodology: SRK, SP, ADS. Writing-review & editing: All authors.

## Data Availability

All data relevant to the study are included in the article. The original MRI data underlying this article cannot be shared publicly due to data protection regulation.

## Additional Information

The authors declare no competing interests related to this manuscript.

