## Supplemental Table for "Strength of spatial correlation between structural brain network connectivity and brain-wide patterns of proto-oncogene and neural network construction gene expression is associated with diffuse glioma survival"

**Supplementary Table 1. Selected Genes.** List of genes for which patterns of transcriptional activity was spatially co-localized with neuroimaging-derived structural connectivity data. Proto-onco genes were selected from the systematic review by Molinaro et al. (1) and neural network genes were selected from the study by Krishna et al. (2).

| **Proto-onco Genes** | **Neural Network Genes** |
| --- | --- |
| *IDH1*  *IDH2*  *TERT*  *ATRX*  *EGFR*  *CDKN2A*  *CDKN2B*  *PTEN*  *TP53*  *NF1*  *MDM2*  *PIK3CA*  *FUBP1*  *NOTCH1* | *SYNPO*  *NTNG1*  *THBS1* |

**Supplementary Table 2.** Drug-gene interactions for the 5 genes that were indicated to be the most important in the survival prediction models (*TERT, ATRX, FUBP1, CDKN2A, SYNPO* and *NOTCH1)*. These genes were entered in the Drug-Gene Interaction Database (DGIdb, v5.0) to explore the potential druggability of genes whose spatial co-localization with gray matter connectivity showed significant association with survival across all predictive models. The Query Score considers the specificity of the gene-drug association, the overlapping associations among genes in the query, and the number of sources supporting the interaction (3).

| **gene** | **drug** | **regulatory approval** | **indication** | **query score** |
| --- | --- | --- | --- | --- |
| NOTCH1 | BRONTICTUZUMAB | Not Approved |  | 22.12 |
| NOTCH1 | RO4929097 | Not Approved |  | 5.53 |
| TERT | CHEMBL:CHEMBL457246 | Not Approved |  | 3.47 |
| TERT | CHEMBL:CHEMBL89977 | Not Approved |  | 3.47 |
| TERT | IMETELSTAT | Not Approved | antineoplastic agent | 3.47 |
| TERT | CHEMBL:CHEMBL520095 | Not Approved |  | 3.47 |
| TERT | TERTOMOTIDE | Not Approved | antineoplastic agent | 3.47 |
| TERT | CHEMBL:CHEMBL144757 | Not Approved |  | 3.47 |
| TERT | SODIUM METAARSENITE | Not Approved |  | 3.47 |
| TERT | CHEMBL:CHEMBL608862 | Not Approved |  | 3.47 |
| TERT | CHEMBL:CHEMBL91163 | Not Approved |  | 3.47 |
| TERT | CHEMBL:CHEMBL89250 | Not Approved |  | 3.47 |
| TERT | CHEMBL:CHEMBL17442 | Not Approved |  | 3.47 |
| TERT | CHEMBL:CHEMBL481161 | Not Approved |  | 3.47 |
| TERT | CHEMBL:CHEMBL190474 | Not Approved |  | 3.47 |
| TERT | CHEMBL:CHEMBL2336666 | Not Approved |  | 3.47 |
| TERT | TELOMESTATIN | Not Approved |  | 3.47 |
| TERT | CHEMBL:CHEMBL1254091 | Not Approved |  | 3.47 |
| TERT | CHEMBL:CHEMBL481160 | Not Approved |  | 3.47 |
| TERT | ISOBONDUCELLIN | Not Approved |  | 3.47 |
| ATRX | TUVUSERTIB | Not Approved |  | 3.28 |
| ATRX | AZ20 | Not Approved |  | 3.28 |
| NOTCH1 | CRENIGACESTAT | Not Approved |  | 2.46 |
| CDKN2A | (10R)-10-METHYL-3-(6-METHYLPYRIDIN-3-YL)-9,10,11,12-TETRAHYDRO-8H-[1,4]DIAZEPINO[5',6':4,5]THIENO[3,2-F]QUINOLIN-8-ONE | Not Approved |  | 2.46 |
| CDKN2A | POLO-LIKE KINASE 1 INHIBITOR GSK461364 | Not Approved |  | 2.46 |
| CDKN2A | HMN-214 | Not Approved |  | 2.46 |
| ATRX | UNC1999 | Not Approved |  | 2.18 |
| TERT | IMETELSTAT SODIUM | Not Approved |  | 1.73 |
| TERT | IODINE I-131 | Approved |  | 1.73 |
| TERT | HLA-A*0201 RESTRICTED TERT(572Y)/TERT(572) PEPTIDES VACCINE VX-001 | Not Approved |  | 1.73 |
| TERT | HTERT-LAMP MRNA-LOADED AUTOLOGOUS DENDRITIC CELL VACCINE GRNVAC1 | Not Approved |  | 1.73 |
| TERT | SURATADENOTUREV | Not Approved |  | 1.73 |
| NOTCH1 | NIROGACESTAT | Not Approved |  | 1.70 |
| NOTCH1 | MK-0752 | Not Approved |  | 1.47 |
| NOTCH1 | ENOTICUMAB | Not Approved |  | 1.47 |
| ATRX | GSK126 | Not Approved |  | 1.31 |
| CDKN2A | PF-00477736 | Not Approved |  | 1.23 |
| ATRX | VE-821 | Not Approved |  | 1.09 |
| ATRX | PARP/TANKYRASE INHIBITOR 2X-121 | Not Approved |  | 1.09 |
| ATRX | CERALASERTIB | Not Approved |  | 1.09 |
| CDKN2A | MILCICLIB | Not Approved |  | 0.98 |
| CDKN2A | PALBOCICLIB | Approved |  | 0.95 |
| ATRX | TAZEMETOSTAT | Approved |  | 0.94 |
| TERT | DENDRITIC CELL VACCINE | Not Approved |  | 0.87 |
| CDKN2A | ABEMACICLIB | Approved |  | 0.77 |
| NOTCH1 | NOTCH SIGNALING PATHWAY INHIBITOR MK0752 | Not Approved |  | 0.74 |
| TERT | OMACETAXINE MEPESUCCINATE | Approved | antineoplastic agent | 0.69 |
| TERT | BDNF | Not Approved |  | 0.69 |
| TERT | ERIBULIN MESYLATE | Approved |  | 0.60 |
| TERT | HTERT VACCINE V934/V935 | Not Approved | antineoplastic agent | 0.58 |
| CDKN2A | RIBOCICLIB | Approved | antineoplastic agent | 0.55 |
| CDKN2A | ULIXERTINIB | Not Approved |  | 0.55 |
| ATRX | TALAZOPARIB | Approved |  | 0.47 |
| TERT | BERBERINE | Not Approved |  | 0.43 |
| CDKN2A | PF-04691502 | Not Approved |  | 0.41 |
| NOTCH1 | HYDROCORTISONE BUTYRATE | Approved |  | 0.40 |
| CDKN2A | TRAMETINIB DIMETHYL SULFOXIDE | Approved | antineoplastic agent | 0.34 |
| CDKN2A | COBIMETINIB | Approved | antineoplastic agent | 0.31 |
| CDKN2A | DABRAFENIB | Approved | antineoplastic agent | 0.29 |
| CDKN2A | SAPANISERTIB | Approved | antineoplastic agent | 0.29 |
| CDKN2A | VEMURAFENIB | Approved | antineoplastic agent | 0.29 |
| ATRX | TOPOTECAN HYDROCHLORIDE | Approved | antineoplastic agent | 0.26 |
| CDKN2A | ALVOCIDIB | Not Approved | antineoplastic agent | 0.26 |
| ATRX | OLAPARIB | Approved |  | 0.25 |
| TERT | RECOMBINANT VASCULAR ENDOTHELIAL GROWTH FACTOR | Not Approved |  | 0.25 |
| ATRX | ADAVOSERTIB | Not Approved |  | 0.23 |
| CDKN2A | BUPARLISIB | Not Approved |  | 0.22 |
| ATRX | TEMOZOLOMIDE | Approved |  | 0.19 |
| CDKN2A | EVEROLIMUS | Approved | immunosuppressant | 0.14 |
| TERT | ARSENIC TRIOXIDE | Approved | antineoplastic agent | 0.13 |
| TERT | 9,10-PHENANTHRENEQUINONE | Not Approved |  | 0.13 |
| CDKN2A | PANITUMUMAB | Approved | antineoplastic agent | 0.13 |
| CDKN2A | LETROZOLE | Approved | antineoplastic agent | 0.13 |
| ATRX | IRINOTECAN HYDROCHLORIDE | Approved | antineoplastic agent | 0.11 |
| TERT | ANTISENSE OLIGONUCLEOTIDES | Not Approved |  | 0.11 |
| CDKN2A | RONICICLIB | Not Approved |  | 0.09 |
| CDKN2A | SIROLIMUS | Approved | for treatment of wet age-related macular degeneration,immunosuppressant | 0.09 |
| TERT | RALOXIFENE HYDROCHLORIDE | Approved | Hormone Replacement Agents | 0.07 |
| ATRX | GEMCITABINE | Approved | antineoplastic agent | 0.07 |
| CDKN2A | GEMCITABINE | Approved | antineoplastic agent | 0.05 |
| ATRX | DOXORUBICIN HYDROCHLORIDE | Approved | antineoplastic agent | 0.05 |
| ATRX | FLUOROURACIL | Approved |  | 0.03 |
| CDKN2A | ILORASERTIB | Not Approved |  | 0.02 |
| ATRX | CISPLATIN | Approved |  | 0.02 |

1.	Molinaro AM, Taylor JW, Wiencke JK, Wrensch MR. Genetic and molecular epidemiology of adult diffuse glioma. Nature Reviews Neurology. 2019;15(7):405-17.

2.	Krishna S, Choudhury A, Keough MB, Seo K, Ni L, Kakaizada S, et al. Glioblastoma remodelling of human neural circuits decreases survival. Nature. 2023;617(7961):599-607.

3.	Freshour SL, Kiwala S, Cotto KC, Coffman AC, McMichael JF, Song JJ, et al. Integration of the Drug-Gene Interaction Database (DGIdb 4.0) with open crowdsource efforts. Nucleic Acids Res. 2021;49(D1):D1144-D51.
